# Capturing the experiences of UK healthcare workers during the COVID-19 pandemic: A structural topic modelling analysis of 7,412 free-text survey responses

**DOI:** 10.1101/2022.06.16.22276487

**Authors:** Danielle Lamb, Liam Wright, Hannah Scott, Bethany Croak, Sam Gnanapragasam, Mary Docherty, Neil Greenberg, Matthew Hotopf, Sharon A.M. Stevelink, Rosalind Raine, Simon Wessely

## Abstract

**Background:** Healthcare workers (HCWs) have provided vital services during the COVID-19 pandemic, but existing research consists of quantitative surveys (lacking in depth or context) or qualitative interviews (with limited generalisability). Structural Topic Modelling (STM) of large-scale free-text survey data offers a way of capturing the perspectives of a wide range of HCWs in their own words about their experiences of the pandemic.

**Methods:** In an online survey distributed to all staff at 18 geographically dispersed NHS Trusts, we asked respondents, “Is there anything else you think we should know about your experiences of the COVID-19 pandemic?”. We used STM on 7,412 responses to identify topics, and thematic analysis on the resultant topics and text excerpts.

**Results:** We identified 33 topics, grouped into two domains, each containing four themes. Our findings emphasise: the deleterious effect of increased workloads, lack of PPE, inconsistent advice/guidance, and lack of autonomy; differing experiences of home working as negative/positive; and the benefits of supportive leadership and peers in ameliorating challenges. Themes varied by demographics and time: discussion of home working decreasing over time, while discussion of workplace challenges increased. Discussion of mental health was lowest between September-November 2020, between the first and second waves of COVID-19 in the UK.

**Discussion:** Our findings represent the most salient experiences of HCWs through the pandemic. STM enabled statistical examination of how the qualitative themes raised differed according to participant characteristics. This relatively underutilised methodology in healthcare research can provide more nuanced, yet generalisable, evidence than that available via surveys or small interview studies, and should be used in future research.

## Introduction

Research examining mental health and wellbeing outcomes for healthcare workers (HCWs) during the COVID-19 pandemic has largely consisted of online surveys. Such methods provide results quickly, involve low burden on participants, and can have relatively large samples. However, many of these studies fail to have sufficiently well-defined samples or response rates to support claims of representativeness^1^. Nonetheless, studies using these methods have found that HCWs report high prevalence of symptoms of psychological distress, anxiety, and depression, with those who are female, nurses, with inadequate access to PPE, and lack of support from superiors or peers tending to report poorer outcomes^2–5^. There is also some evidence that key workers (including HCWs) report higher levels of depression and anxiety symptoms than non-key workers^6^.

It is widely accepted that self-report surveys, while providing interesting and sometimes useful snapshots of the prevalence of mental health symptoms, often miss deeper and more detailed insights about why people may be struggling, and how they could be better supported. In addition, surveys tend to over-estimate the prevalence of mental health disorders^7^. Qualitative research can offer depth and nuance about the factors associated with poor mental health and what sorts of support processes may be beneficial, but typically is conducted using one-to-one interviews or small focus groups. This drastically limits the number of participants and variety of perspectives that are feasible to include. While generalisability is not typically the goal of qualitative research^8^, from a policy-making and organisational perspective it is desirable to hear from a large and diverse sample in order to ensure systems and support are set up in ways that benefit as many of the workforce as possible.

One way to gain valuable, open-ended insights from a much greater volume of participants than traditional one-to-one interviews, is to provide respondents to quantitative surveys with free-text boxes to report openly on specific questions posed. While there are limitations, e.g. the inability for an interviewer to ask follow up questions or probe more deeply, the requirement that participants are literate (and computer literate/have computer access, where the survey is online), there are also advantages in terms of confidentiality (leading, potentially, to more honest reporting), and the ease and convenience of data collection for participants and researchers^9^. Where the quantity of data collected is prohibitive to manual coding, e.g. where thousands of responses have been collected, computerised text mining methods provide a way to help analyse responses.

This study used structural topic modelling^10^, a text mining technique, on free-text responses to an online survey investigating healthcare workers’ experience of the COVID-19 pandemic between April 2020 and January 2021. Structural topic modelling (STM) enables the extraction of themes from free-text responses and the quantitative summary and analysis of these themes alongside other participant data such as demographics, using regression models. This method has been used previously in health science^11^, but remains relatively underutilised in this area of research. The aim of this study was to use STM to analyse responses from a broad question about HCW experiences of the pandemic, to allow HCWs to identify what was most salient and important to them.

## Methods

### Ethics

Ethical approval for the study was granted by the Health Research Authority (reference: 20/HRA/210, IRAS: 282686) and local Trust Research and Development approval. Cohort data are collected via Qualtrics online survey software, pseudonymised and held on secure university servers. Participants are aware that they can withdraw from the study at any time, and there is signposting to support services if participants feel they need it. Only those consenting to be contacted about further research will be invited to participate in further components.

### Data collection

The data used in this analysis comes from the NHS CHECK study (for more information about the wider study, see ^12^, a longitudinal cohort study of the mental health and wellbeing of HCWs during the COVID-19 pandemic. All staff members in 18 participating NHS Trusts were invited to complete an online survey once between April 2020 and January 2021. At the end of the survey was a free-text box where participants were asked, “Is there anything else you think we should know about your experiences of the COVID-19 pandemic?”. The responses to this question form the data used in this analysis.

### Data analysis

We performed STM, implemented with the stm R package^13^ to extract topics from responses. We performed STM using unigrams (i.e. single words). STM treats documents as a probabilistic mixture of topics, and topics as a probabilistic mixture of words. STM is a “bag of words” approach that uses correlations between word frequencies within documents to define topics. An advantage of STM is that it allows for inclusion of covariates in the estimation model, so the estimated proportion of a text devoted to a topic can differ according to document metadata (e.g., participant characteristics). We included participant’s age (modelled with natural splines with 3 degrees of freedom to account for potential non-linearities in the association: ^14^, date of response (natural splines, degrees of freedom 3), sex, marital status (married, co-habiting, single, divorced, widowed), ethnicity (White, Mixed Race, Black, Asian, Other) and NHS trust (18 categories) in our models, each collected at baseline. There was only a small amount of item missingness (6.1% for age; 2.2% for sex; 2.1% for marital status; 1.9% for ethnicity; 3.8% for role), so we used complete case data.

Prior to performing the analysis, we cleaned free-text response using an iterative process. We excluded responses containing fewer than five words and removed words which appeared in fewer than five responses. We also removed common “stop” words (“the”, “and”, “I”, etc.) from the analysis. Spelling mistakes were identified with the Hunspell spellchecker^15^, amended manually if they had five or more occurrences and replaced using the Hunspell suggested word function if the number of occurrences was fewer than five. Where the algorithm provided multiple suggestions, the word with the highest frequency across responses was used. To reduce data sparsity, in the structural topic models, we used word stemming using the Porter^16^ algorithm (e.g., help, helping and helped become help). Data cleaning was carried out in R version 3.6.3^17^.

We ran STM models, which included between two to 50 topics and selected the final models based on visual inspection of the residuals and lower bound statistics of the model solutions. After selecting a final model (with 35 topics), we carried out two analyses, a thematic analysis of the topics and qualitative text excerpts, and a regression analysis of participant characteristics and generated themes. In line with previous work using this methodology^18^, for the thematic analysis, we generated short descriptive titles for the 35 topics, firstly individually, and then via collaborative discussion. We dropped one topic due to incoherence between the exemplar texts, and merged two topics due to similarity. The remaining 33 topics were grouped, again, initially individually, and then in an agreed thematic structure via a collaborative process involving interpretation of the content of the topics. Descriptive summaries were written for each theme and subtheme, and the thematic analysis proceeded via an iterative process of reading and re-reading exemplar texts. We identified quotes that captured the essence of each of the themes generated, and reflected on meanings and interpretations of these in terms of the overarching narrative of the themes and data, and in terms of our own perspectives. While the generation of themes was supported by STM, rather than initial familiarisation with every text excerpt (which, while usual in reflexive thematic analysis, was not possible with over 7,000 responses), the subsequent stages of meaning-making and discussion of our own positionality was in keeping with recent guidance regarding reflexive thematic analysis^19^. The researcher team comprised women and men, researchers and clinicians, and diversity in terms of age and ethnic background. We undertook extensive discussion of our subjective interpretations of the data, and how these may have been influenced by our own experiences and perspectives.

### Patient and public involvement

Frontline NHS staff proposed this research, and we tested the proposal’s acceptability and approach with a small informal reference group of front-line staff (psychologists, managers, intensivists and trainee psychiatrists) and refined it accordingly. We have developed an advisory group of NHS staff (clinical, managerial, auxiliary, students), who meet online and contribute virtually to provide input on methods, recruitment strategy, communications, and interpretation of findings.

## Results

### Descriptive Statistics

Sample descriptive statistics are displayed in Table 1. There were 7,412 participants (35% of total sample) who provided a valid free-text response. The distribution of responses according to age, sex, marital status, ethnicity and NHS trust was similar among those who provided a free-text response and those who did not.

**Table 1.** Descriptive statistics

| Variable | Categories | All Participants | Missing % | Valid Free-Text Response |
| --- | --- | --- | --- | --- |
| Sample Size | - | 21,190 | - | 7,412 (35%) |
| Age | - | M:42.8 (SD: 12.0) | 6.1% | M: 44.91 (SD: 11.5) |
| Sex | Male | 3,832 (18.5%) | 2.2% | 1,272 (17.2%) |
|  | Female | 16,896 (81.5%) |  | 6,140 (82.8%) |
| Marital Status | Married | 10,117 (48.8%) | 2.1% | 3,797 (51.2%) |
|  | Co-habiting | 5,152 (24.8%) |  | 1,691 (22.8%) |
|  | Single | 3,884 (18.7%) |  | 1,253 (16.9%) |
|  | Divorced | 1,401 (6.8%) |  | 587 (7.9%) |
|  | Widowed | 193 (0.9%) |  | 84 (1.1%) |
| Ethnic Group | White | 17,819 (85.7%) | 1.9% | 6,304 (85.1%) |
|  | Mixed ethnic or racial group | 524 (2.5%) |  | 219 (3%) |
|  | Black | 880 (4.2%) |  | 367 (5%) |
|  | Asian | 1,416 (6.8%) |  | 463 (6.3%) |
|  | Other ethnic or racial group | 154 (0.7%) |  | 59 (0.8%) |
| Role | Doctor | 1,566 (7.7%) | 3.8% | 565 (7.7%) |
|  | Nurse | 5,260 (25.8%) |  | 1,968 (27.0%) |
|  | Other clinical | 6,263 (30.7%) |  | 2,356 (32.3%) |
|  | Non-clinical | 7,304 (35.8%) |  | 2,412 (33.0%) |

### Thematic Analysis

From the 33 topics, two overarching domains were identified, personal (45.8% of text), and professional (52.9% of text), each containing four themes (see Table 2), although many themes contained elements of both the personal and the professional, and were often interlinked.

**Table 2.** Theme table

| **Personal** |  | **Professional** |  |
| --- | --- | --- | --- |
| **Social impact** | **Restrictions and rules** | **Home working** | **Workplace culture** |
| Social isolation<br>3% | Adherence/risk<br>3.3% | Home working/shielding<br>8.5% | Teamwork/support<br>4.4% |
| Contribution<br>2.7% | Rules<br>1.6% | Home schooling/<br>childcare<br>4.1% | Leadership/ managerial<br>support<br>3.5% |
| Bereavement<br>2.5% | Trust in Government<br>3.1% | Physical impacts<br>1.7% | Media/politicians<br>1.7% |
| Caring responsibilities<br>1.8% |  |  | Support/help<br>1.7% |
| **Mental health** | **Physical health** | **Workplace challenges** | **New roles** |
| Anxiety/adaptation<br>3% | Vaccination<br>3.5% | Workload/burnout<br>4.5% | Changes to work<br>pattern/role/<br>redeployment<br>4% |
| Mental health impact<br>2.8% | Testing<br>3.5% | Personal Protective<br>Equipment (PPE)<br>3.2% | Starting/ending roles<br>3.9% |
| Impact on life<br>2.8% | COVID-19 symptoms<br>1.8% | Challenges to clinical<br>practice<br>2.6% | Job roles/students<br>3% |
| Anger/personal<br>challenges<br>2.5% | Fear of infection<br>2.2% | Satisfaction with<br>employment<br>1.9% |  |
| Personal mental health<br>1.3% | Shielding<br>2.7% | Stress<br>1.5% |  |
|  | Risk/comorbidity<br>1.8% |  |  |

The personal domain contained themes related to physical health (15.4% of text), mental health (12.4% of text), social impact (10.0% of text) and restrictions and rules (8.0% of text). The professional domain contained themes related to home working (17.1%) of text), workplace challenges (13.7% of text), workplace culture (11.3% of text), and new roles (10.9% of text). See Figure 1.

**Figure 1.**
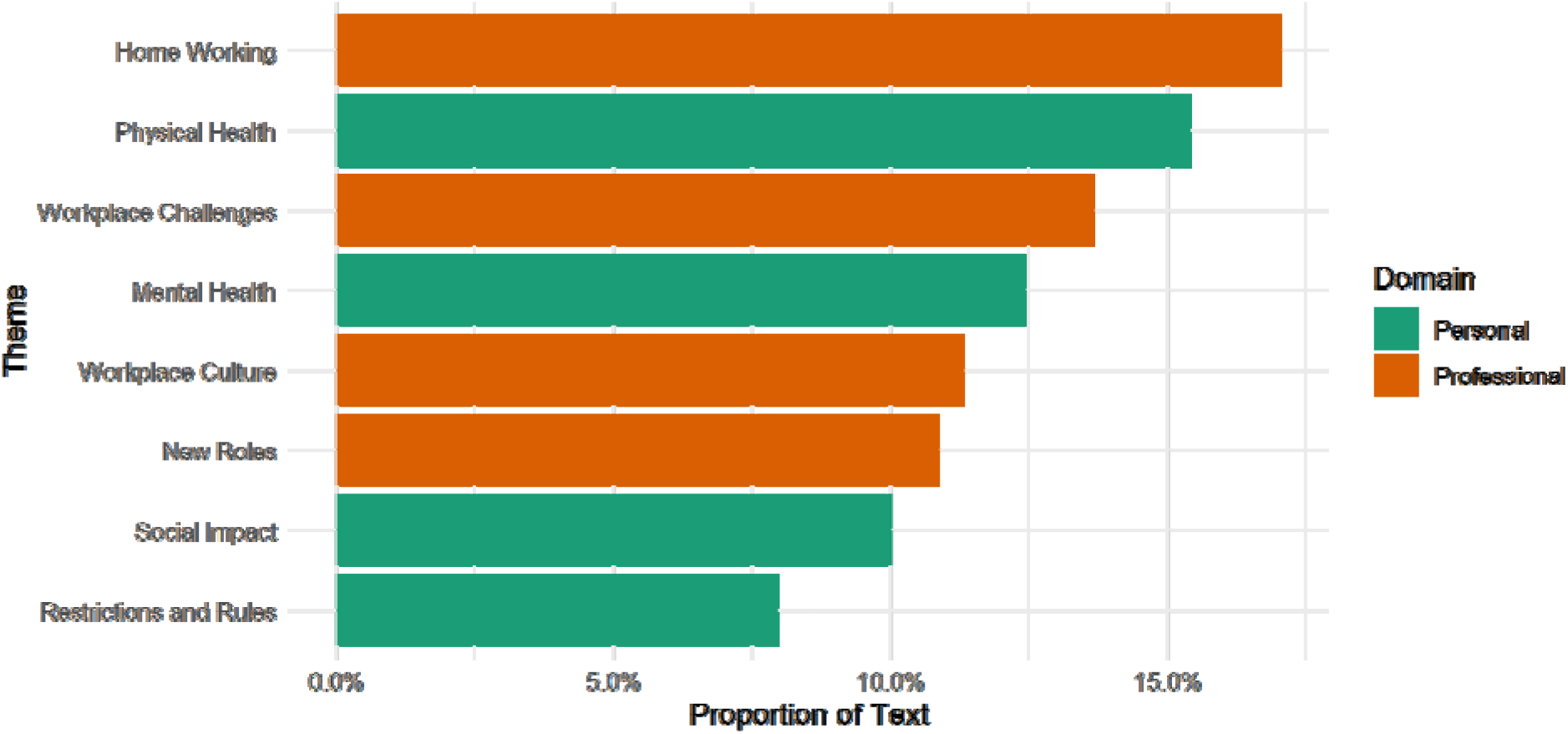
Proportion of text devoted to each theme.

After each quote used to illustrate the themes below is the role of the participant, grouped by job role (doctors, nurses, Allied Health Professionals (AHPs), and non-clinical staff). Sub-themes are indicated by bold text.

#### Personal

##### Social impact

Many participants discussed how the pandemic and subsequent lockdown restrictions prevented them from fulfilling their personal **caring responsibilities**, which was upsetting for them and their relatives:

> *“Having an elderly bed bound mum, who has carers visiting her, and it was extremely difficult for me and her when I couldn’t visit her during lockdown.” (Non-clinical staff)*

Some participants faced challenges due to schools and nurseries being shut, for example, *“Hard to balance work and childcare especially as single parent.” (Doctor)*.

**Social isolation** was identified as challenging: *“Not being able to see family members or friends quite upsetting” (Non-clinical staff)*. Respondents reported the struggles of not seeing family and friends for long periods of time due to not living locally, and several respondents even described the breakdown of their relationships due to the time spent apart, for example, *“This has contributed to the end of my marriage.”* (AHP). These issues negatively their affected mental health, as identified in the theme below.

Some respondents reported that being isolated from colleagues by working from home also impacted on them personally, feeling as though they were making less of a **contribution** than those in hospitals: *“Working at home can be isolating sometimes and normal work can seem less relevant making you feel less useful than clinical colleagues.” (Non-clinical staff)*.

Participants described various experiences of **bereavement** (mostly deaths of relatives) during the pandemic, some directly caused by COVID-19. Some detailed how the emotional toll of these bereavements has been exacerbated by the social isolation the restrictions have caused and not being able to be comforted in person by loved ones, for example, *“Due to bereavement and family illness I have been significantly impacted due to not being able to see family.” (AHP)*.

##### Pandemic restrictions and rules

For some, the response at the national political level was felt to be poor, with a lack of **trust in Government**. The frustration was particularly related to the pace of the response, considered as slow by some, as well as advice being conflicting, with mixed messages: *“Poor and conflicting information from Government” (Non-clinical staff)*. Others felt there was a disconnect between the practice undertaken at their organisation and Government guidelines, for example, *“Conflicting information from management vs Government information is stressful.” (Non-clinical staff)*.

A number of participants expressed considerable frustration with seeing individuals not adhering to **rules** such as social distancing. Some felt such actions were *“selfish and inconsiderate” (AHP)*, and reported that this made them feel angry:

> *“I have been angry to how people are allowed to flout or twist the rules imposed by the Government during this pandemic. I am also angry that the Government do not do more to reduce the cases.” (AHP)*.

Others expressed difficulty with **adherence** to rules, particularly in a healthcare setting, given the stringent nature of the rules, as well as their constant changes and related confusion – *“It’s difficult sticking to the regulations within healthcare settings and it’s all overwhelming” (Nurse)*.

##### Physical health

Many respondents described their experience of **COVID-19 symptoms**, such as loss of taste, headaches, fatigue, and cough. Some commented on the longevity of their symptoms:

> *“Mild initial infection has led to long-standing problems with breathlessness, dry cough, hoarse voice, concentration problems, balance issues, neurological problems, unexplained rashes, skin lesions and crushing fatigue at times.” (Non-clinical staff)*.

Participants reported not being able to get antigen *testing* despite symptoms, which fuelled further frustration and distrust in government, but some discussed the positives of participation in **vaccination** trials:

*“I’m also participating in the Oxford Vaccine Trial and am appreciating regular swab tests from this. It is very nice to have feedback that at the time of each test I know my likely COVID status.” (Doctor)*.

Some felt that those who were **shielding** or had recognised **risk/comorbidity** factors were being looked down upon by colleagues due to their inability to carry out their normal duties, and these feelings were damaging to morale and team cohesion.

> *“COVID related bullying is happening and no one (even managers) know how to deal with it. I loved my job before COVID; now we are divided. People with underlying health concerns are treated as being slackers. Laughed at for being concerned and there is an age split between young and older.” (AHP)*.

In addition, many respondents expressed **fear of infection** with COVID-19. Participants commented that this fear was coupled with the worry that they would then pass on the infection to their household: *“Fear of getting the virus and giving it to my husband or other member of my family. Fear of dying of it myself and leaving my husband.” (Nurse)*. Certain factors seemed to exacerbate this concern such as their family member having an underlying health condition or being in a high risk group: *“Very worried about having BAME [Black and minority ethnicity] partner and bringing the virus back to them.” (Nurse)*.

##### Mental health

Staff reported feeling overwhelmed, experiencing **anxiety**, and having sleep problems with vivid dreams and nightmares, for example, *“I have been woken most nights with nightmares and/or vivid dreams which does not normally happen.” (Nurse)*, with uncertainty being a key contributing factor:

> *“Uncertainty, changes happening so frequently with no consultation to staff on the floor until things are actually happening. Uncertainty of the future for our dept.” (AHP)*.

Participants working in the mental health services noted that they felt that their services were *“completely forgotten” (AHP)* relative to physical health services. The significant work-based pressures in mental health service were reported as impacting negatively on their own mental health, and therefore their ability to care for others:

> *“The number of referrals for patients with serious mental health difficulties have increased significantly causing a huge strain on the service and my own mental health.” (AHP)*.

Positively, some participants highlighted the support from their organisation and senior leaders such as Chief Executives. This was felt to be protective, which is echoed in the ‘Workplace culture’ theme below:

> *“The positive messages from our chief executive have been key to maintaining mental health against the onslaught of negativity and speculation from the media.” (Non-clinical staff)*.

With regards to the wider **impact on life**, a number of participants experienced loneliness and isolation during the pandemic, as noted above. This was particularly related to separation from family, friends, romantic partners or being single - *“Being a single person has led to increased loneliness and self-doubt about all aspects of my life” (Non-clinical staff)*.

On the other hand, some participants noted personal benefits during the pandemic in being able to spend more time with family and developing a supportive social support network:

> *“We as a family have actually been happier during this period once we’d got over the infection. The pace of life has slowed down and given us more quality time together. For us this has been the silver lining to this sad situation.” (Non-clinical staff)*.

Participants expressed frustration with being labelled a hero, and wished for better working conditions, pay, and support instead:

> *“The whole ‘heroes’ thing was awful, and the clapping mawkish and largely insincere. Hero statues means we don’t need normal basic human needs. It will also be forgotten immediately and we will be bullied into doing more to catch up without recognition, reward, and with threats and bullying. Working in the NHS will be worse than before, despite the superficial societal praise.” (Doctor)*.

For some HCWs, these challenges resulted in diagnosis of **personal mental health** conditions and the need for treatment from health services, for example:

> *“I worked in ITU [Intensive Care Unit] during the first surge and have since been diagnosed with PTSD, depression, anxiety and binge eating disorder. Started on anti-depressants and awaiting intense CBT therapy for the PTSD.” (AHP)*.

In addition, some staff members with pre-existing mental health conditions faced exacerbation during the pandemic, *“I am Bipolar and had an episode of depression at this time, largely precipitated by having to do things ‘virtually’”* (Non-clinical staff). This was related to work and social stressors, as well as disruption in access to support services.

#### Professional

##### Workplace challenges

A major challenge reported by staff was the pressure of **workload**. Respondents described the stress of increasing workloads and understaffing, in part due to staff sickness with COVID-19, but also due to increased bed capacity or caseloads without commensurate increases in staffing:

> *Any stress I experience comes about due to my staff having very high caseloads and increasing referrals with staff not having caseload space to take these people on and be able to do a good job for them. (Non-clinical staff)*.

Staff report a ‘vicious cycle’ of increased workloads leading to **stress** and **burnout**, which in turn lead to staff off sick, with fewer to pick up the resulting work, and difficulties in retaining staff. For example:

> *We have been massively understaffed throughout COVID due to staff sickness - mostly down to stress rather than having to isolate. This has impacted on the stress levels of the remaining staff who are firefighting trying to stay afloat. (Non-clinical staff)*.

These issues were apparent for clinical and non-clinical staff, and are summed up by one respondent’s comment, that, *“The pressure on staff is unsustainable.” (Nurse)*.

Respondents reported the adverse effects of shortages of **PPE**, as well as feeling that where PPE was available it was not of high quality or sufficient to protect workers:

> *The lack of PPE equipment has been terrifying. The visors appropriate to operate on are being kept hidden because there aren’t enough and the PPE visors (not on a surgical mask) don’t protect from upward blood splashes. It is becoming dangerous to do our job, even more so than usual due to failings of ordering and providing correct equipment. (Doctor)*.

Even for those with adequate access to PPE there were problems, with some reporting that equipment did not fit properly, *“There are no FFP3 masks that fit me so I feel unsafe.” (AHP)*, and a lack of training: *“I do not think the training for PPE (donning and discarding) was adequate, especially for community staff.” (Doctor)*.

There were multiple **challenges to clinical practice**, with the unpredictability exacerbating existing stress, and potentially reducing the quality of patient care:

> *“The main stressor for me is the unpredictability at work. You never know if you are going to be moved or if the ward you’re on is going to swing from green to red, or back. Resulting in massive patient movement and extra work.” (Nurse)*.

Lack of consultation left staff feeling undervalued, and changes in the types of work being undertaken also caused stress, as staff felt unprepared and not able to provide adequate levels of care:

> *“We were redeployed in the first wave without consultation as a testing unit….we were an older peoples medicine ward and we were without warning taking patients from A+E who were quite unwell and outside of our normal expertise… we have had constant trouble sourcing equipment for unwell patients. Again, we were not consulted … which felt like the Trust saying they did not value us.” (Nurse)*.

Although some staff expressed **satisfaction** with aspects of their work, for example, *“Been really happy that I have been able to continue offering a service to our patients” (AHP)*, others reported deep dissatisfaction and intentions to leave their roles: *“It’s made me decide to retire.” (Non-clinical staff)*. Some respondents felt that management were indecisive and unsupportive:

> *“Management seem unable to decide how we should offer psychotherapy to clients, whether we should go into work or not. there is a feeling we are being ‘scrutinised’ rather than supported.” (AHP)*.

Staff reported lack of access to employment rights, wellbeing resources, and equipment to allow homeworking, and some experienced frustration and ethical dilemmas at not being able to offer high quality care:

> *“Many of us in the NHS have been unable to deliver services to our patients because of services being stopped. For me this has led to feelings of guilt, frustration and moral injury.” (AHP)*.

##### Home working

Participants often mentioned **working from home** due to **shielding:** *“I’m shielding so I’ve been working at home and felt very disconnected from work.” (Non-clinical staff)*. For some, working from home has been helpful in providing control and autonomy, whereas for others, the blurred boundaries with other family members and lack of in-person support from colleagues has been challenging:

> *“I find working from home has helped A LOT with my mood, life and with work. I feel more in control and when I have bad mental health days being at home is the best thing for me and I don’t need to take a day off. I can still work and get my tasks done which has a positive effect on my mood.” (Non-clinical staff)*.
>
> *“The impact from working at home in when you work in mental health is negative. Conversations about family abuse, rape, incest and violence now happen in my home. At times whilst my children are in the next room. These conversations have happened in my bedroom. The boundaries between home and work were extremely important for mental health. The little bits of informal supervision you get in the office and ways you can debrief after a traumatic session are all lessened by working from home.” (AHP)*.

An issue often raised by participants was managing work alongside **home-schooling**, and the challenge of these competing demands:

*“Home-schooling primary school age children is not compatible with a full-time job and the provisions offered by schools were very varied.” (Non-clinical staff)*.

Participants reported that an additional challenge to homeworking was the **physical health impact**, commonly noting musculoskeletal difficulties resulting from not having the correct seating/desk/computer equipment to work in a comfortable position, and the eye strain/headaches caused by long hours looking at screens:

> *“Working from home has been extremely difficult. Not the correct equipment. I’ve had to self-refer to physiotherapy for back, shoulder and neck problems.” (AHP)*.

##### New roles during the pandemic

A number of participants described **starting or ending roles** during the pandemic unrelated to redeployment. A small number of participants told us that they had been made redundant, that they had difficulty finding a new role, or that beginning their new role had presented additional challenges given remote working/workload pressure:

> *“Started my role in February just before so this has affected on how I have adapted the way I work before really even getting to grips with the role.” (Nurse)*.

Others referenced temporary **redeployment** and the deep anxiety around this, in particular the fear of providing inadequate care to patients:

> *“I found being sent to adult ITU extremely difficult, it made me anxious and terrified to come to work […] I am a PICU [Pediatric Intensive Care Unit] nurse not an adult nurse and was left alone to take full responsibility for extremely ill COVID adult. I felt out of my depth, scared and it has made me and my colleagues to worried about a second peak and being sent again.” (Nurse)*.

For others however, redeployment was a positive experience:

> *“I felt the time before redeployment was more stressful than actually being redeployed. My time at the Nightingale was very positive because everyone was very supportive.” (Nurse)*.

This suggests that the experience was very individual and often related to the team, colleagues and other work-based support systems, and this is echoed in the ‘Workplace culture’ theme below.

##### Workplace culture

In contrast to some of the challenges outlined above, many respondents reported how well supported they felt by colleagues, highlighting the importance of **teamwork**. For example, staff found it helpful to continue working with existing colleagues, *“Being able to work with my same colleagues has been really helpful and I feel has made us stronger as a team.” (Nurse)*. Others also noted the fact that well-supported staff function better, *“It has shown that teamwork, support and compassion are needed, when that is in place people function better.” (Non-clinical staff)*.

Examples were given of the practical steps team leaders have put in place to support staff, for example:

> *“As a single parent with a young child I have been really well supported by my team leader and rest of my team…My team leader set up a buddy system and we each had a buddy who we contacted each day for a video chat, this was such a help and I am extremely grateful to have such supportive colleagues in the NHS.” (AHP)*.

While those with good **leadership/managerial support** coped well, others reported poor relationships, and the negative impacts of these. Senior management in some areas were criticised for lack of communication:

> *“I did not feel supported by senior managers. My direct manager was great but was not supported herself. The trust did a terrible job of communicating and appeared panicked and out of control.” (Nurse)*.

Honesty from senior managers, as well as regular, clear communication, and addressing problems in practical, visible ways were all appreciated by staff, summarised neatly by this participant, *“Compassionate leadership, genuine compassionate leadership is key.” (Non-clinical staff)*.

The **support** provided by Trusts was perceived as key to helping staff to feel valued, and to perform well. Free parking was mentioned repeatedly as being one-way Trusts can support staff, for example, *“The Trust has been very supportive to staff - allowing free parking has been monumental in my wellbeing during this time.” (AHP)*. However, pressure for services to see as many patients as possible was seen as unhelpful, especially given most staff value providing quality care: *“Pushes on flow rather than getting things right has been a feature pushed through by Director of Operations which has been unhelpful and quite frankly – frustrating.” (Non-clinical staff)*.

Beyond the workplace, the **political** leadership of the country has impacted on staff wellbeing, for example, one participant was particularly blunt about their feelings:

> *“The single biggest factor in my mental health around coronavirus has been the shambolic handling of the situation by the Government. I have no faith or trust in their leadership, and that is a toxic situation in a pandemic such as this.” (Doctor)*.

Similarly, the role the **media** played was also criticised, with a sense of resignation about whether those in positions of responsibility will be held to account:

> *“The fear mongering media and politicians have not come out of this looking good. Stats have been manipulated and lies told. Will it all ever come out and accountability made*…*I doubt it.” (Non-clinical staff)*.

### Regression Analyses

The results of the regression models are displayed in Supplementary Figures S1-S3. There were some differences in themes raised by age (Figure S1). Notably, older participants were more likely to discuss challenges related to physical health and to home working, while younger participants were more likely to discuss new roles. There were also differences according to the date free-text responses were recorded (Figure S2). As may be expected, discussion of home working decreased across the pandemic, while discussion of workplace challenges increased. Discussion of mental health was lowest between September-November 2020, the period between the first and second waves of COVID-19 in the UK.

Differences according to sex, marital status and ethnic group were also observed (Figure S3). Notably, women were more likely to discuss home working, new roles and social impact, and were less likely to discuss restrictions and rules. There were also differences by NHS trust, most notably regarding positive and negative experiences of home working and physical health (results not shown for privacy reasons).

## Discussion

### Summary of results

Responses from the 7,412 participants fell broadly into two domains, personal and professional. though there was overlap in the content of several themes, with important connections between home working, isolation, and HCW mental health. Furthermore, we found evidence that demographic factors affected the way the respondents reported their experiences of the pandemic, and that experiences also varied depending on the nature of national picture of the pandemic at the time the data was collected. This paper presents qualitative data from the largest UK survey of HCW wellbeing, with demographics broadly representative of the wider NHS workforce ^20^. It is likely that participants reported the most salient experiences for them, and thus the responses provide insight into important factors that may not have been picked up by other research using pre-defined survey questions or fixed interview schedules.

One, perhaps unsurprising, finding was that the personal challenges most commonly reported by HCWs overlapped considerably with those faced by the general population (e.g. bereavement, social isolation, caring responsibilities). However, we also identified specific workplace challenges for HCWs that are less relevant for many other types of workers (e.g. facing significant risk of infection, redeployment), and these in turn impacted home life. That said, our sample was not homogenous, and variations between participants’ experiences within the workplace suggest that guidance around infection control, vulnerable staff, and redeployment differed between Trusts and teams. This also demonstrates important differences in personal preferences for ways of working, and underlines the challenges for Trusts in offering support that is appropriate for all members of staff.

### Results in relation to previous research

Some of our findings echo the results of previous quantitative work in this area especially in that increased workloads were a key concern for HCWs ^2^. Many of our participants reported increased workloads, with a vicious cycle of this leading to burnout and sickness absence, which in turn led to further increased workloads for those remaining; this has been noted in other qualitative research ^21,22^. As our regression analyses showed, discussion of workplace challenges increased over time, indicating the growing importance of these as the pandemic progressed. However, concerns about heavy workloads would not have been solely due to the pandemic, though it has clearly exacerbated them. The 2018 NHS Staff Survey found that 40% of HCWs reported work-related stress, burnout, and dissatisfaction due to increased workload, which was caused by staff shortages ^23^. In our findings and in other research, inconsistency and uncertainty at work are repeatedly mentioned as stressful, and causes of negative wellbeing ^24–27^, as were redeployment and changes to working patterns. In contrast, and also found in quantitative studies, peer support and effective leadership help to reduce negative outcomes ^21,26,28^.

Lack of PPE was a concern for our participants, and has also been found to negatively impact of employee mental health in non-healthcare settings ^29^. A number of qualitative studies have found similar results, that access to appropriate PPE is an important factor for HCWs’ mental and physical wellbeing ^26,27,30^, though our evidence suggests a temporal trend, with more concerns about this in the early months of the pandemic. There is evidence that access to PPE has been an issue internationally ^28^ and in previous pandemics ^22^. Lack of PPE, or access to inadequate PPE, as well as changing guidance about its use, have impacted HCW trust in leaders and politicians, and there have been particular concerns about differential access to PPE, e.g. by women and Black, Asian and minority ethnic HCWs ^30^.

Fear of infecting loved ones was frequently reported as an important source of stress. This has been reported previously ^21,22^, as have the challenges of balancing personal and professional stressors. For example, home working was experienced positively by some, providing time to slow down and connect with family, but others reported this to have been a negative experience, with social isolation, home schooling while trying to work, and physical health impacts all issues of concern ^31,32^. These findings were somewhat gendered, with women more likely to discuss home working than men, and this echoes previous qualitative research looking at female HCWs’ experiences of the pandemic ^33^.

### Implications for policy and practice

Whilst the provision of clear and appropriate guidance on working practices is needed, employers of HCWs need to provide flexibility to cater to differing social needs and preferences. We identified the highly detrimental effects of uncertainty on staff wellbeing, and we suggest that proactive, clear communication from senior managers, and Government, is likely to positively impact on the mental health of staff.

We also identified that peer support and compassionate leaders were viewed as mitigating stress, and may prevent the onset of formal mental health problems. These views correlate well with findings from quantitative studies that have examined other trauma-prone occupational groups, which show that supportive junior leadership and effective peer support are associated with better employee mental health ^34^. Training for managers may help to reduce the highly variable support participants reported in this study, and could be particularly beneficial given the fact that HCWs seem to want immediate support from those around them, rather than from well-being counsellors on a help line. Brief training for managers to improve their confidence to speak to their staff about mental health has been shown to be effective ^35^. Alongside strong employee social bonds, our results also suggest Trusts should invest in ensuring that staff are able to access both wellbeing support measures (e.g. wellbeing spaces) as well as access to professional support from occupational health teams and, where needed, primary and secondary care services.

One aspect of our findings that has been consistently echoed in other research during the COVID-19 pandemic, and of previous pandemics, is the necessity for HCWs to be provided with appropriate and adequate PPE. While this is an issue that also needs to be addressed at the political level, healthcare leaders should be aware of the urgency in advocating for their staff on this issue.

### Strengths and limitations

Firstly, the most important strength of this study is that the data analysed are drawn from the largest UK study of HCWs, with a relatively representative sample from 18 English Trusts. Secondly, and relatedly, the STM methods used enabled us to include the views of a much larger sample than is typically possible in qualitative work, and allowed us to statistically examine how the topics raised differed according to participant characteristics. Thirdly, the open phrasing of the question allowed us to capture issues that are important to HCWs, and as such, we have generated a more holistic picture of the key issues than more tightly focussed qualitative studies can offer. This includes the interplay between the personal and the professional, such as the influences of personal circumstance, organisational culture and support, and the role of Government and wider society on respondents’ perceptions. Fourthly, the anonymity offered by the online data collection method means some participants may have felt able to be more honest about challenging topics than they would have done in a face-to-face or virtual interview.

However, the online nature of the survey highlights the first limitation of this research, that technical capability and access were required to participate, as well as English language skills, which may have excluded the most vulnerable. Secondly, as the free-text question from which the data was drawn was not compulsory, and was in addition to survey that covered a range of wellbeing topics, we might have been more likely to capture the views of participants with stronger or more extreme experiences. Thirdly, while the STM enabled inclusion of a large sample, the modelling is no more objective than the researchers interpreting findings; some topics covered a number of different areas meaning that researchers had to use their own judgement about which comments most clearly captured the main point of a topic, and about how to group the topics into meaningful themes. To mitigate this as far as possible, four researchers collaborated in an iterative process of individual and group interpretation. Fourthly, the data were collected over a relatively long time-period (April 2020-January 2021) so do not capture the mood of a specific era of the pandemic, though conversely this does mean we were able to map the themes identified over time.

### Implications for future research

The method used in this study clearly demonstrates the benefits of large-scale qualitative data collection. Such methods can capture factors that survey designers might have missed as important to participants, and can add context, depth, and nuance to quantitative results. STM has not been widely used in health research to date, despite the frequent inclusion of free-text responses in large surveys. We advocate the use of these methods. Future research could helpfully address how to mitigate some of the limitations of these methods, such as how to capture the views of those with limited access to online surveys, or cultural differences in interpretations.

## Conclusion

This analysis of free-text data, from 7,421 participants in the UK’s largest survey of HCWs, provides in-depth, nuanced evidence of the most salient factors affecting them during the COVID-19 pandemic. In line with previous research, increased workload, lack of PPE, uncertainty, and inconsistency in messaging and advice from leaders were experienced as negatively impacting worker wellbeing, while support from peers and managers helped staff to cope with personal and professional stressors.

## Data Availability

The free-text data used in this study cannot be made publicly available due to stipulations set out by the ethics committee. The code used in the analysis is available at https://osf.io/4d8tf/.

## Author contributions

SW, NG, MH, RR, and SS are chief investigators of the study, and contributed to manuscript drafts and approved the final draft. DL is a co-investigator of the study and led the write-up of the manuscript. LW performed the STM and regression analyses and led the write up of these sections of the manuscript. DL, HS, BC, and SG performed the thematic analysis and wrote these sections of the manuscript. MD is a collaborator of the study and contributed to manuscript drafts.

## Acknowledgments

We wish to acknowledge the National Institute of Health Research (NIHR) Applied Research Collaboration (ARC) National NHS and Social Care Workforce Group, with the following ARCs: East Midlands, East of England, South West Peninsula, South London, West, North West Coast, Yorkshire and Humber, and North East and North Cumbria. They enabled the set-up of the national network of participating hospital sites and aided the research team to recruit effectively during the COVID-19 pandemic.

The NHS CHECK consortium includes the following site leads: Sean Cross, Amy Dewar, Chris Dickens, Frances Farnworth, Adam Gordon, Charles Goss, Jessica Harvey, Nusrat Husain, Peter Jones, Damien Longson, Richard Morriss, Jesus Perez, Mark Pietroni, Ian Smith, Tayyeb Tahir, Peter Trigwell, Jeremy Turner, Julian Walker, Scott Weich, Ashley Wilkie.

The NHS CHECK consortium includes the following co-investigators and collaborators: Peter Aitken, Anthony David, Sarah Dorrington, Rosie Duncan, Cerisse Gunasinghe, Stephani Hatch, Daniel Leightley, Ira Madan, Isabel McMullen, Martin Parsons, Paul Moran, Dominic Murphy, Catherine Polling, Alexandra Pollitt, Danai Serfioti, Chloe Simela, Charlotte Wilson Jones.

